# A New, Simple Projection Model for COVID-19 Pandemic

**DOI:** 10.1101/2020.03.21.20039867

**Authors:** Jian Lu

## Abstract

**Background:** With the worldwide outbreak of COVID-19, an accurate model to predict how the coronavirus pandemic will evolve in individual countries becomes important and urgent. Our goal is to provide a prediction model to help policy makers in different countries address the epidemic outbreak and adjust the control policies to contain the spread of the severe acute respiratory syndrome coronavirus 2 (SARS-Cov-2) more effectively.

**Methods:** Unlike the classic public health and virus propagation models, this new projection model takes both government intervention and public response into account to generate reliable projections of the outbreak 10 days to 2 weeks in advance. This method is an observation based projection similar than the classic Moore’s Law in miroelectronics. The Moore’s law is not based on any physics law and yet has anticipated the development of microelectronics for decades. This work is an empirical relation to decribe the evolution of epidemic to pandemic situations in different countries. The country was selected as an observation unit because the regulation and political decision is an national decision for numerous measures such as the implementation of social distancing, the quarantine of suspected cases, and the closing of borders to achieve territorial containment.

**Findings:** This model has been successfully applied to predict the evolution of pendemic situation in China. Then the model was also validated by the South Korean data. With a reduction of cases calculated as reduction coefficient of the increase rate of daily cases Rc = 2% per day, we observed a very efficient policy with a strict systematic control in both China and South Korea. For the moment, the Canada, USA, Australia may have difficulties to limit the fast evolution of the epidemic. With a Rc<0.5%, it’s particularly important for the USA to consider escalating the control measures because the affected cases can reach more than one million very soon.

**Interpertation:** Due to the difference of national disciplines and historical culture, the national policy may be implemented and observed with different efficiency. The starting point where the government decided to apply total containment can also play a key role for the evolution of the pendemic situation. The model will allow each national government of the nations still affected by the pandemic to project the situation for the coming 10 to 14 days. It’s very important for the deployment of national and international efforts to stop the pandemic situation.

**Funding:** National Key R&D Program of China (Ministry of Science & Technology (MOST, China))

## 1. Introduction

The classic epidemic models are mainly based on the capacities of the virus to propagate in an environment without protection. The variables include disease incubation period, speed and strength of viral propagation while introducing some assumptions related to the slowdown barriers. However, reality could be a lot more complicated. For instance, COVID-19’s incubation period varies from 1 to 14 days. What’s more, to stop epidemic spread and reduce infections to lower levels is a concerted effort of both the government and the people. The strategies of dealing with the outbreak, the strength of government execution, how people react and respond to the measures are critical to the number of infections.

In addition, facing such a great uncertainty, it is impossible to think of the future in terms of a single result but rather a range of possibilities. To project the possibility of a variety of outcomes, a model derived from real propagation data that can summarize the possibilities and describe their relative likelihood will say a lot – but not everything - about the likely future. Otherwise, the results are likely to generate a huge scatter of errors.

Making use of primary data at the initial stage of the outbreak, right after the drastic containment measure imposed in different countries and regions, this new projection model is an integrative multi-parameters model that evaluates the effectiveness of the government interventions while forecasting the spread of virus. As mentioned above, such effectiveness is affected by two other major factors: public response (including how much awareness they have, the level of their cooperation) as well as efficiency of healthcare system (including the capacity, advancement, and effectiveness).

## 2. Model and Method

In any given country, the outbreak usually starts slow, followed by a rapid-growing unstable period. The length of this unstable period depends on the performance of initial defence and control measures in each country and how swift government intervention is. For COVID-19, we found the rate of increase in the number of infections was around 1.3 during the unstable period in China. All the datas used in this work are from the Reference 1.

China declared a level 1 health emergency on January 29 evening. On January 30, transport suspension in Hubei and Beijing began. This new model was first built with the data of infected cases in Mainland China starting from February 1.

While charting the number of infected cases since February 1, we project different reduction coefficient of daily cases increase rate (*R*_*c*_) which is a combined indicator of the effectiveness of government intervention, public response, and healthcare system. Five *R*_*c*_*(s)* give out five scenarios and five projectionn curves based on the formula below:

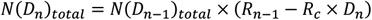

where *D*_*n*_ is the number of days starting from the first simulation day to day *n*;

*N(D*_*n*_)_*total*_ is the total number of infected cases till day *n*;

*R*_*n-1*_ is the case increase rate prior to day *n*;

*R*_*c*_ is the reduction coefficient of daily cases increase rate.

The value of *R*_*c*_ varies according to the effectiveness of government intervention, the awareness and response of the public as well as the efficiency of healthcare system. Therefore, it is different among countries. For China, we used *R*_*c*_ as 0.03, 0.02, 0.015, 0.0125, 0.01.

When *R*_*n*−1_ − *R*_*c*_ × *D*_*n*_ <1%, the increase percentage will be fixed at 1% per day for the coming 5 days and 0.5% for another 5 days to predict the total number of infected cases.

### 3. Prediction Results and Analysis

#### 3.1 Greater China

The figure 1 shows the prediction figure and the real confirmed cases every day in China. The spread of COVID-19 was stable with the numbers of infections increase along with the *R*_*c*_ curve of 1.5% before February 12. The leapt on February 12 was due to a change of China’s methodology of calculation by including people diagnosed via CT scans as well as via testing kits in Hubei.

**Figure 1.**
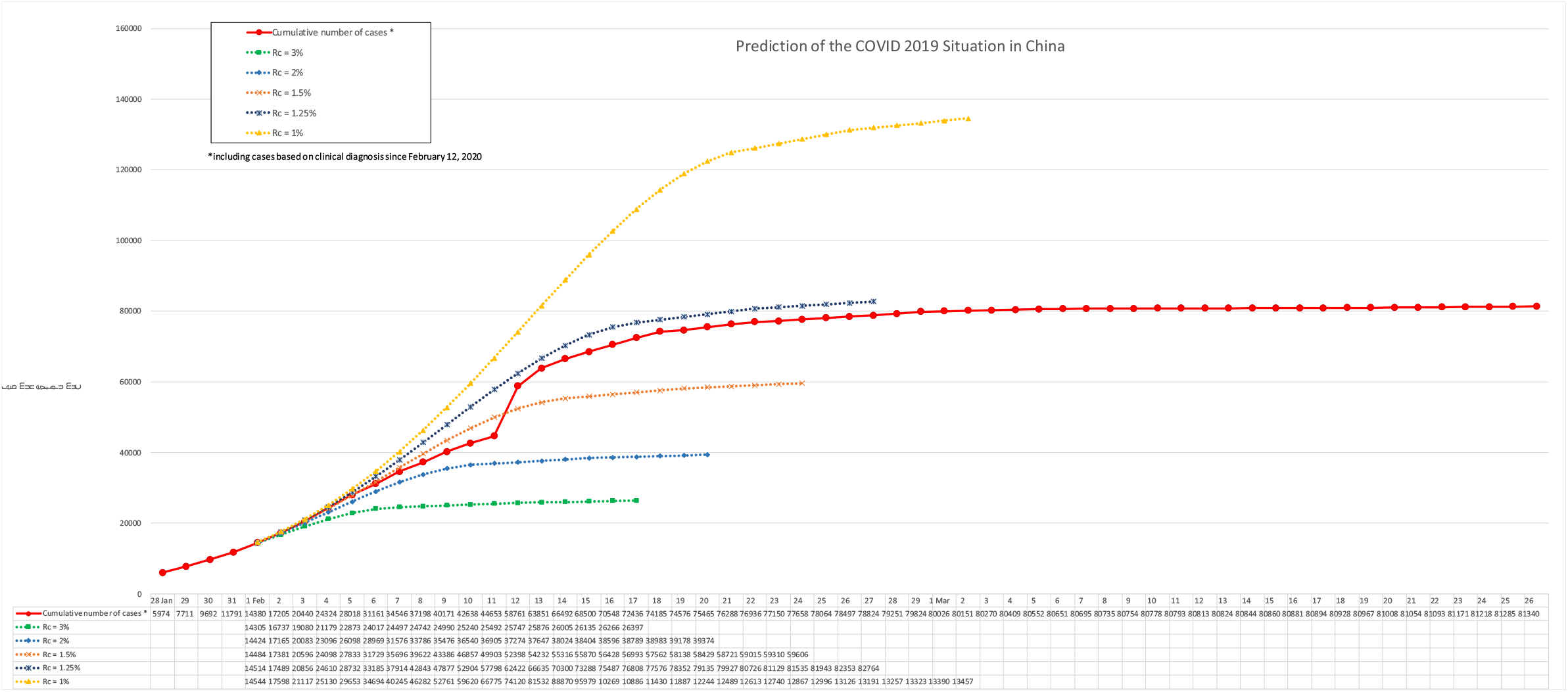
Prediction and real evolution in China.

Afterwards, the projection of *R*_*c*_ 1.25% curve gave out the total number of 82764 on March 27, which was close to the actual total infections of 81285.

#### 3.2 South Korea

The figure 2 shows the prediction figure and the real confirmed cases every day. The number of infected cases went along with the *R*_*c*_ 2% curve. Afterwards, the projection of *R*_*c*_ =2% curve gave out the total number of 8323 on March 19, which was close to the actual infections of 8565 for the same day.

**Figure 2.**
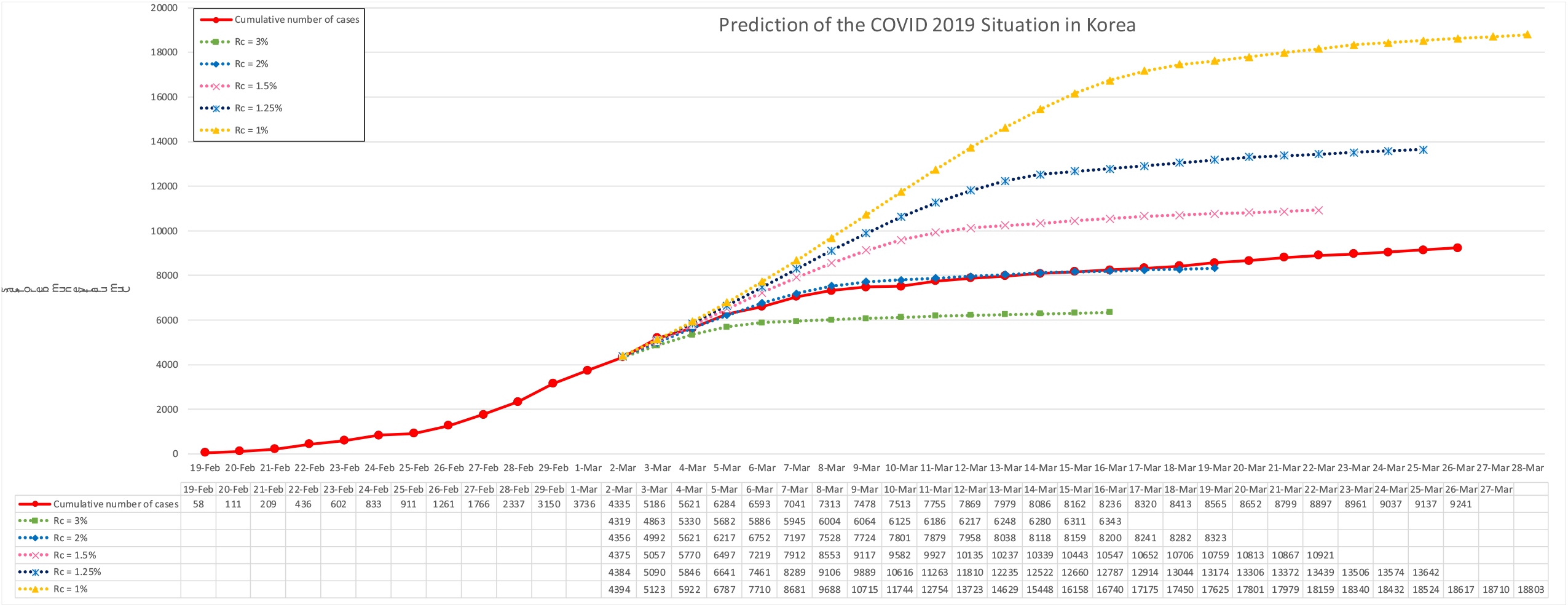
Prediction and real evolution in South Korea.

#### 3.3 Iran

The figure 3 shows the prediction figure and the real confirmed cases every day in Iran. Iran has a *R*_*c*_ =1.5% before 22 March. But it’s now moving progressively to a dangesous zone showing a new phase of evlution.

**Figure 3.**
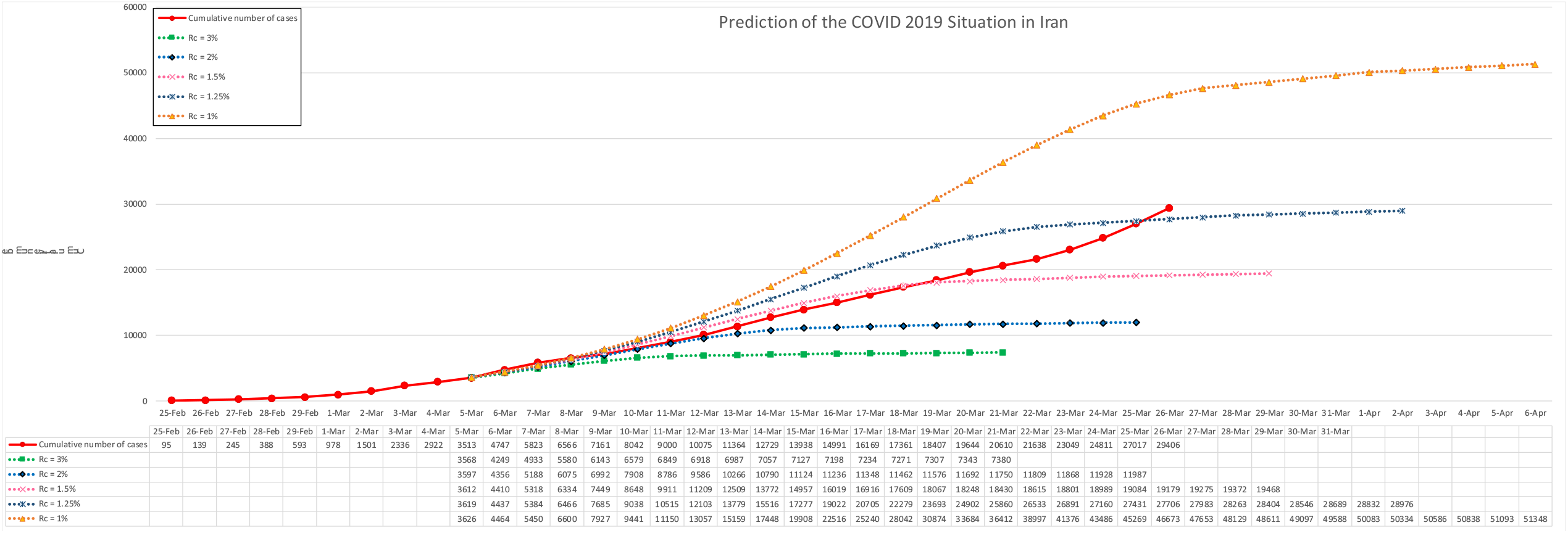
Prediction and real evolution in Iran.

#### 3.4 Italy

The figure 4 shows the prediction figure and the real confirmed cases every day in Italy. Italy’s *R*_*c*_ is close to 1.25%, indicating a total number of cases could reach 56971 on 5^th^ April 2020. Italy’s *R*_*c*_ is now moving from 1.25% to 1%, showing an important deterioration of the disease control which may generate more than 100,000 cases.

**Figure 4.**
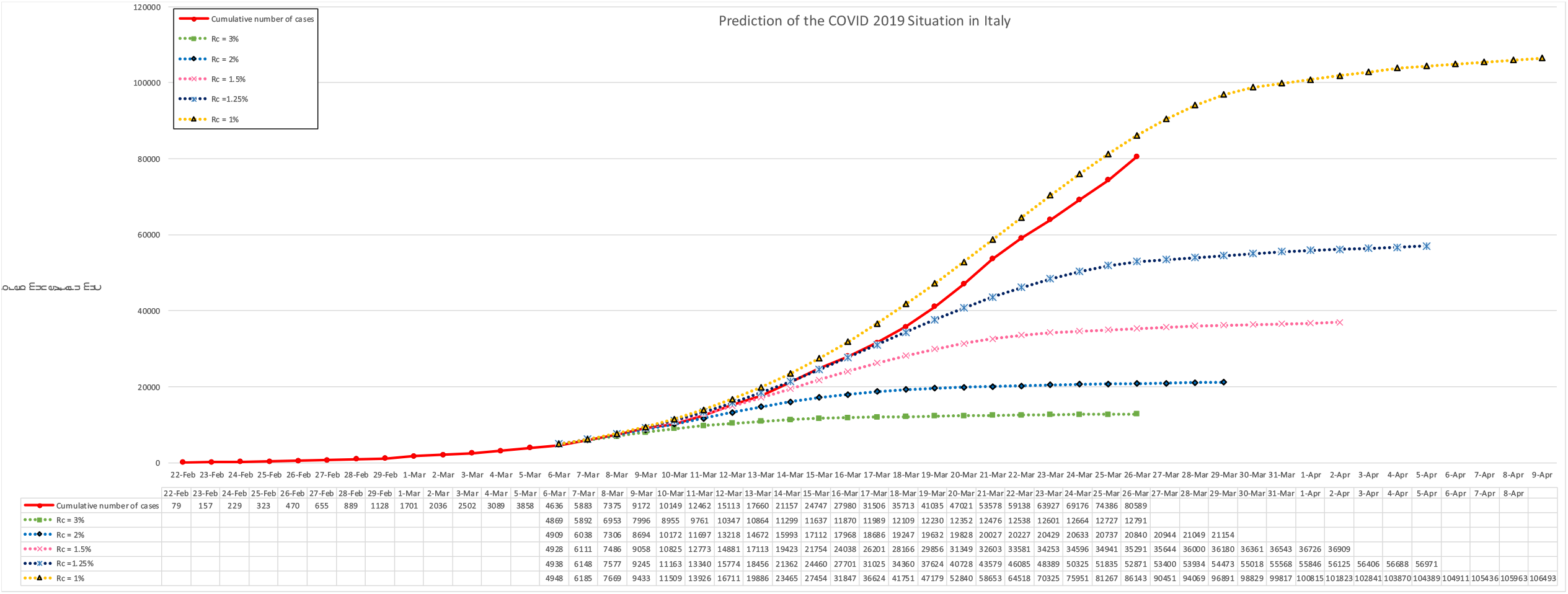
Prediction and real evolution in Italy.

#### 3.5 France

The figure 5 shows the prediction figure and the real confirmed cases every day. France’s *R*_*c*_ is also around 1.25%.

**Figure 5.**
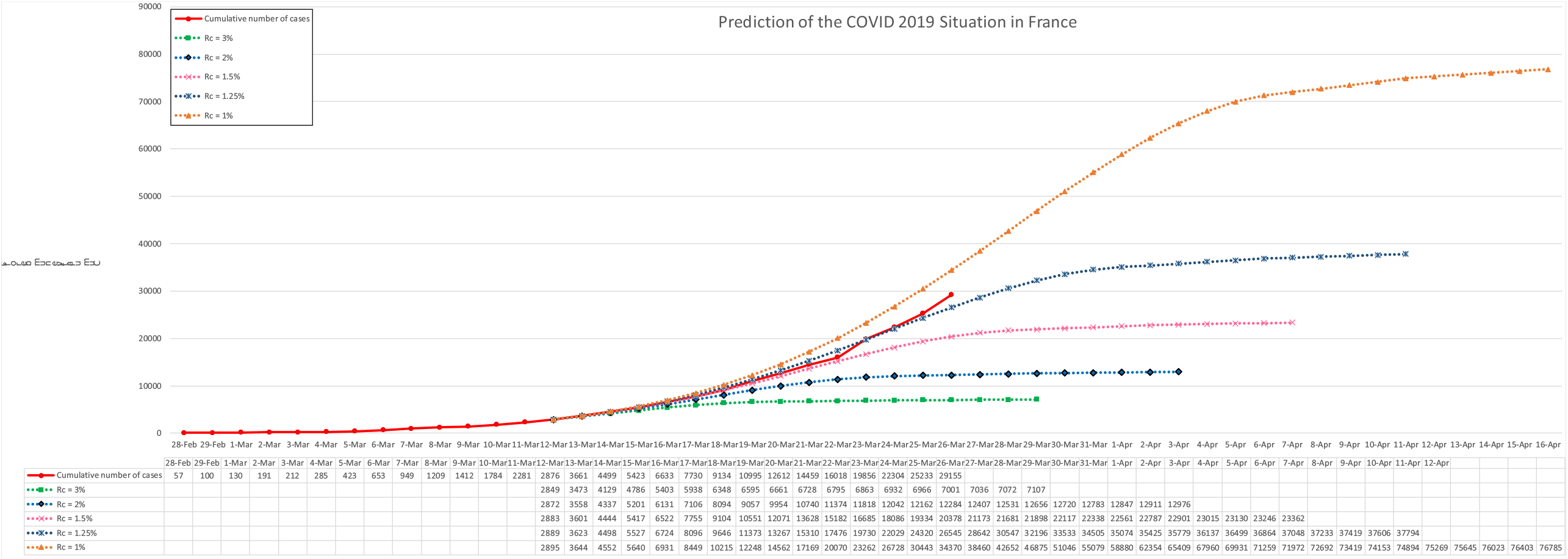
Prediction and real evolution in France.

#### 3.6 Germany

The figure 6 shows the prediction figure and the real confirmed cases every day. Germany’s *R*_*c*_ moved from 3% to 2%, showing a deterioration of the disease control but still under control.

**Figure 6.**
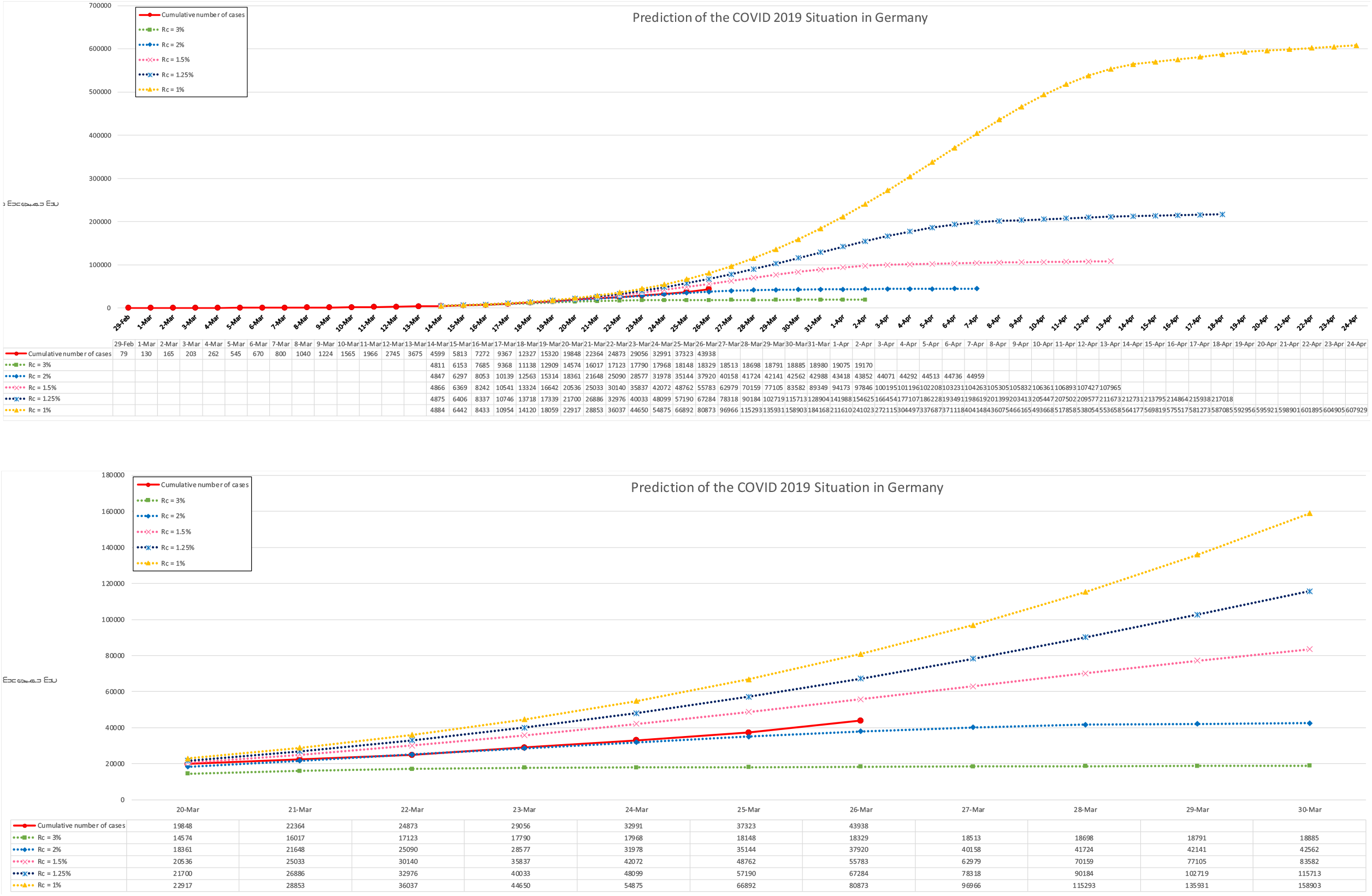
Prediction and real evolution in Germany.

#### 3.7 USA

The figure 7 shows the prediction figure and the real confirmed cases every day in USA. USA’s *R*_*c*_ is less than 0.5 % by mid-March, indicating that there could be at least a million infections (by when?) if the situation does not improve.

**Figure 7.**
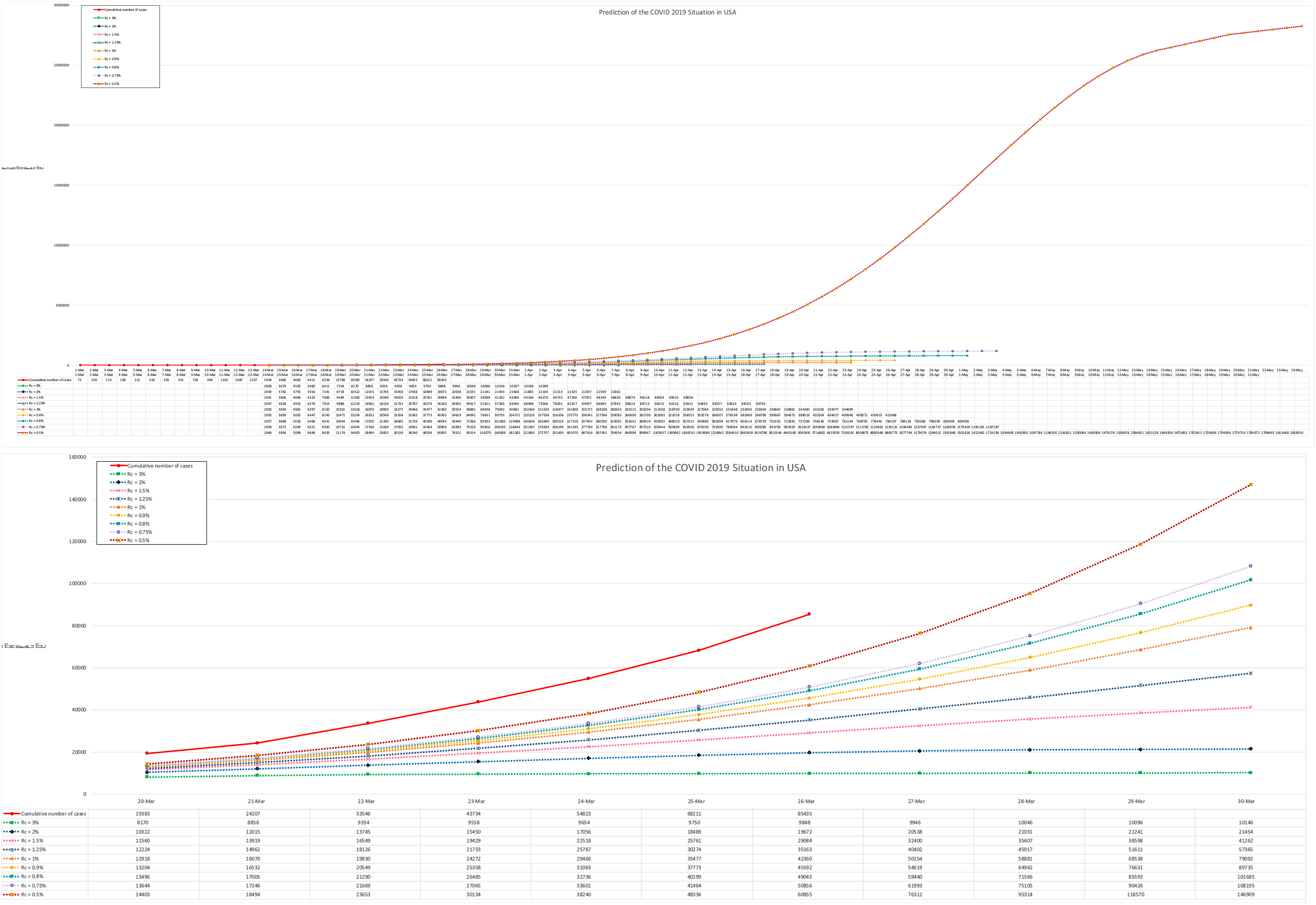
Prediction and real evolution in USA.

#### 3.8 Canada

The figure 8 shows the prediction figure and the real cases in Canada. A Rc less than 0.5% showing that the situation is not yet under control.

**Figure 8.**
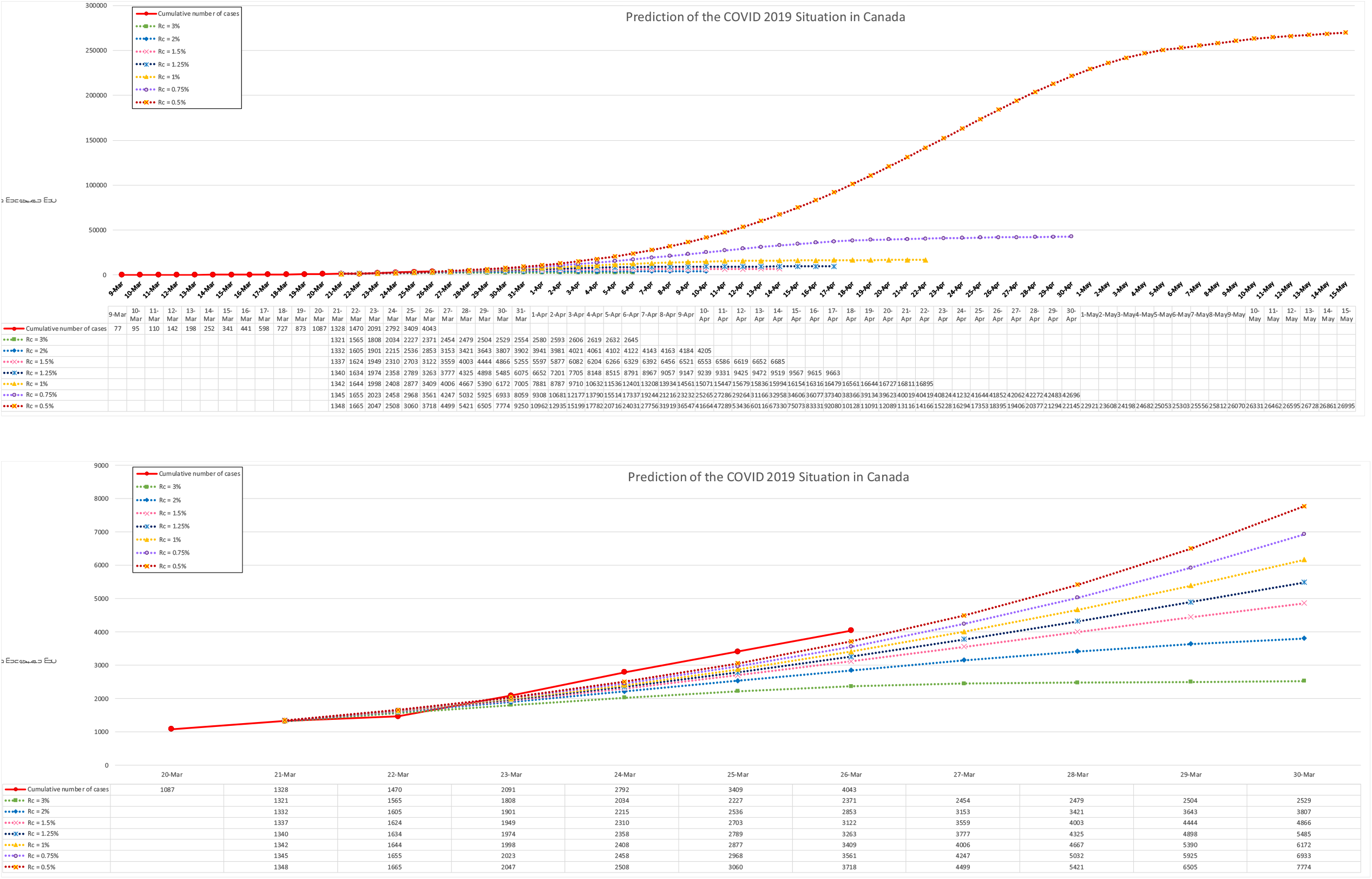
Prediction and real evolution in Canada.

#### 3.9 Australia

The figure 9 shows the prediction figure and the real cases in Canada. A Rc less than 0.5% showing that the situation is not yet uneder control. The total number of affected cases can reach a number more than 100,000.

**Figure 9.**
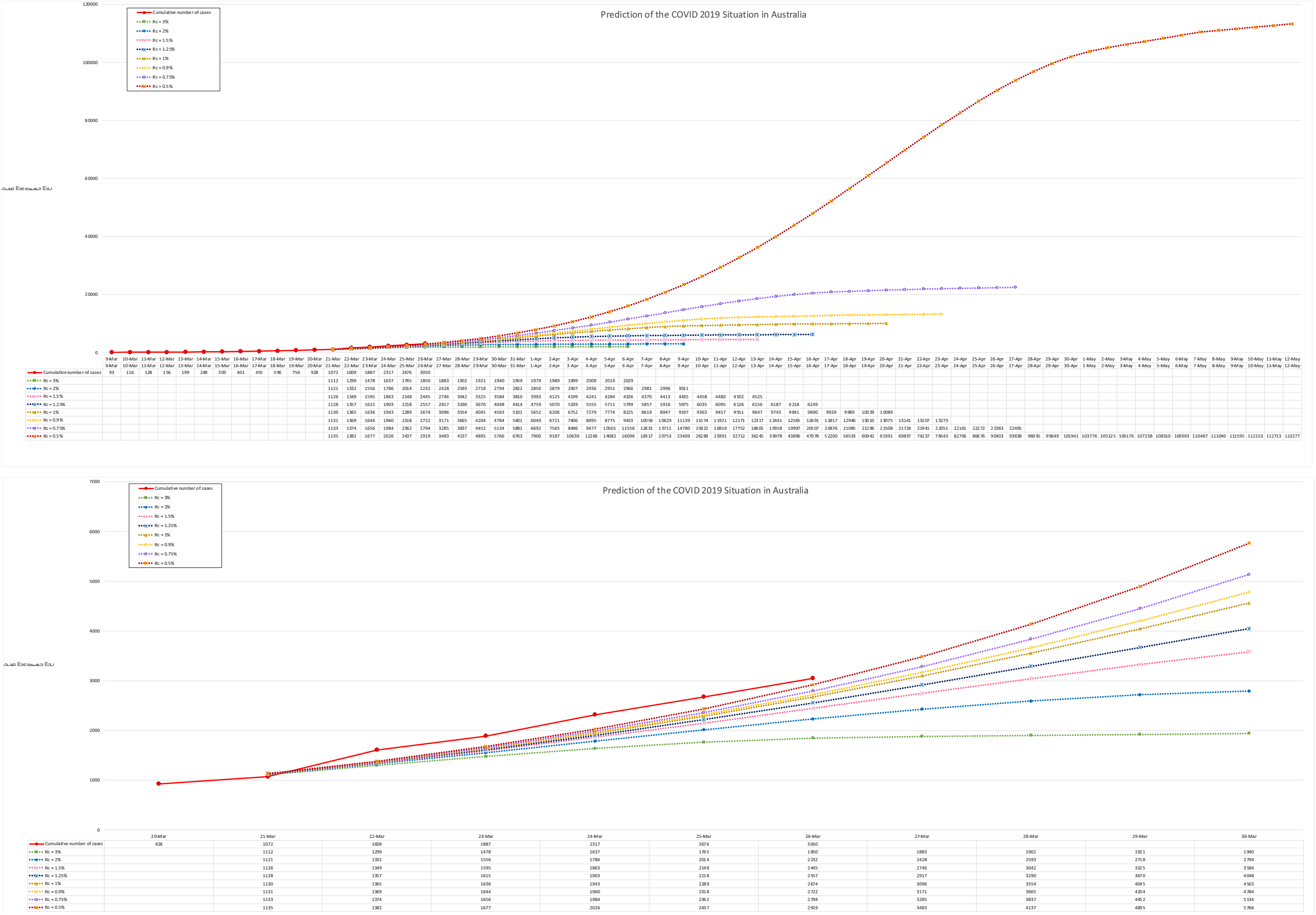
Prediction and real evolution in Australia.

The figure 10 shows the the prediction figure and the real cases in UK. Although the strategy of the British government has changed from herd immunity to a more realistic policy, with a Rc=1.25% the results shows that the country is still under the risk to have more than 50000 cases by 18 April 2020 if we observe closely the trends of the evolution of confirmed cases.

**Figure 10.**
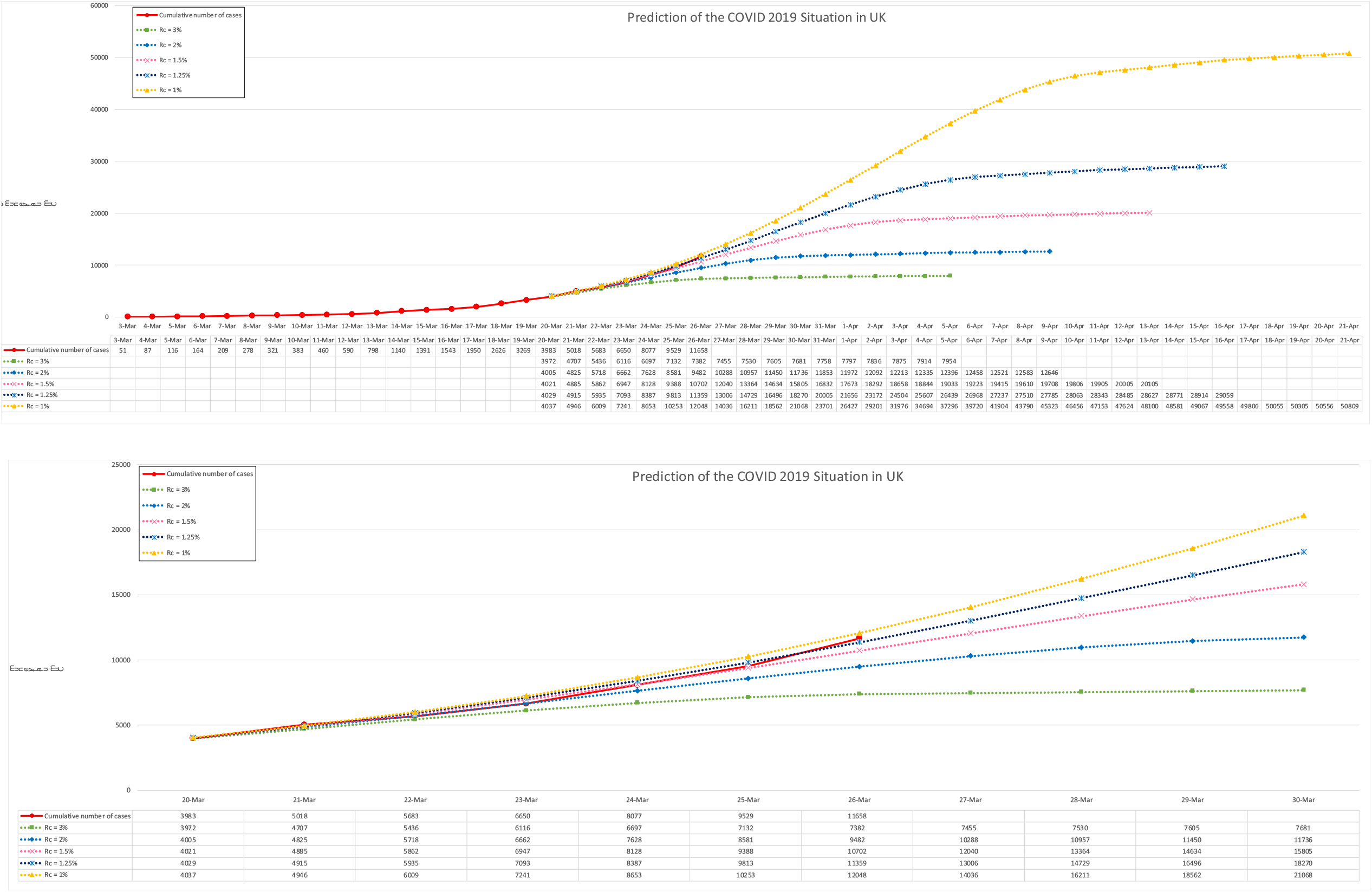
Prediction and real evolution in UK.

Based on above simulation, we can draw a world map (figure 11) to observe the worldwide evolution of the pendemic situation. This is a simple but accurate method appreciate the world under the pendimic situation.

**Figure 11.**
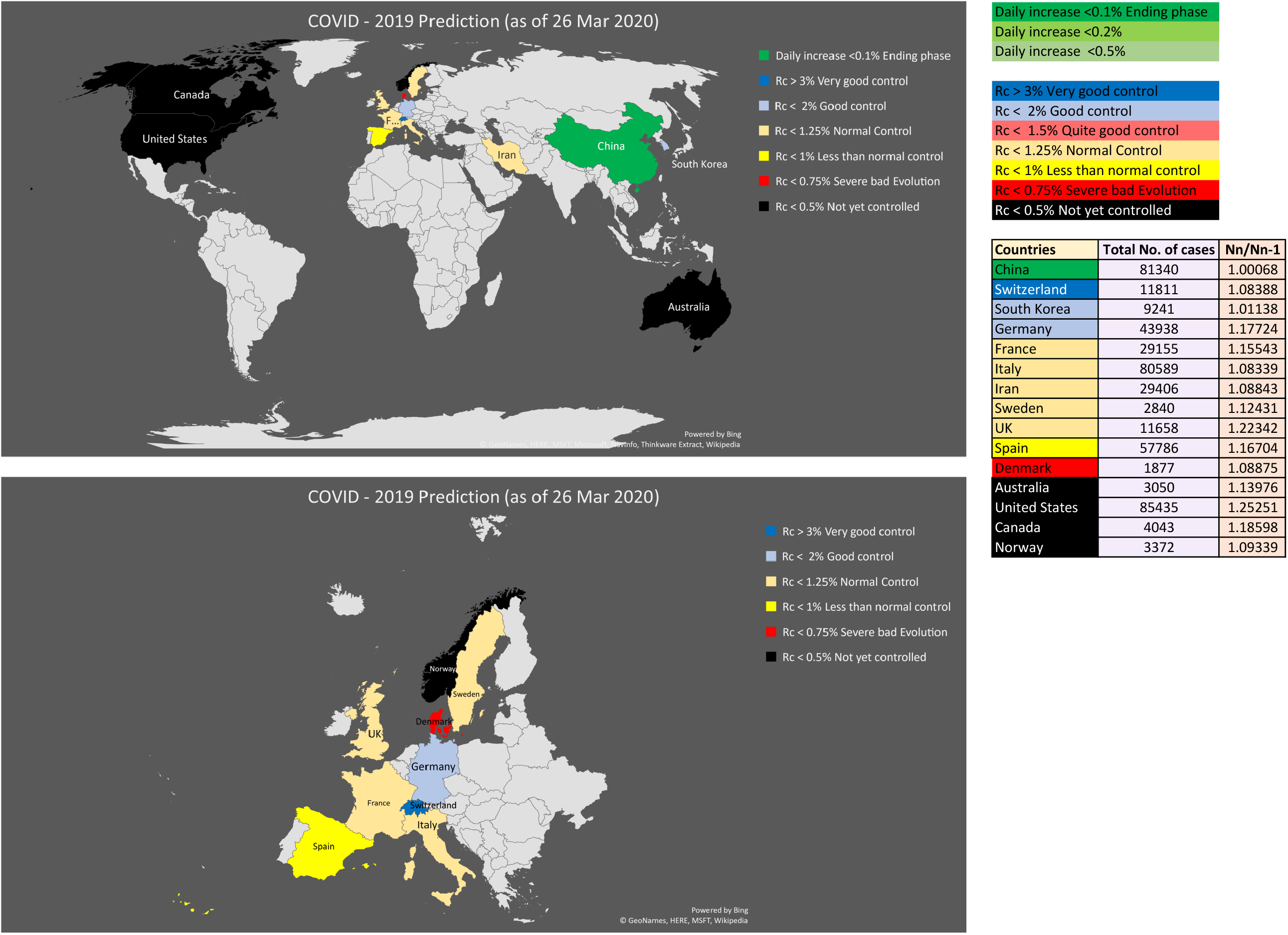
Prediction and real evolution world map.

## Conclusion

The new, simple projection model had successfully applied to the epidemic situation in Mainland China and offered a decent, reliable prediction of the number of infected people for the coming two to four weeks. The model outlines which outcome is most likely and what other outcomes also have a good chance of occurring, how broad the range of possible outcomes can be. It offers an idea of the evolution of the infection situation and the projection model is applicable to other countries like South Korea, Iran, Italy, France, USA, and Germany.

This projection model aims to achieve two major tasks of epidemic control: one is to assess the effectiveness of government intervention, the level of cooperation from the public and the efficiency of healthcare system; second is to track and give a decent prediction of the growth and number of infections. To eradicate a pandemic, both information is critical for policy makers to outline strategies and make adjustment to measures to curb the spread of infection, minimizing the social and economic impact to one’s country and to the world.

## Data Availability

All data of confirmed COVID-19 cases are from the website:https://www.worldometers.info/coronavirus/

## Acknowledgments

J. Lu gratefully acknowledges that the work was supported by the National Key R&D Program of China (Project No. 2017YFA0204403)

